# Machine learning models for tuberculous pleural effusion diagnosis in Africa setting

**DOI:** 10.1101/2025.01.18.25320778

**Authors:** Eric Walter Pefura-Yone, Adamou Dodo Balkissou, Laurent-Mireille Endale-Mangamba, Jodie Bane, Massongo Massongo, Marie Elisabeth Ngah Komo, Virginie Poka-Mayap, Alain Kuaban, Abdou Wouliyou Nsounfon, Djenabou Amadou, Arnaud Laurel Ntyo’o’ Nkoumou, Paul Ledoux Tendap-Ndam, Marie Josiane Ntsama-Essomba, Marie Christine Ekongolo, Christian Mbobara-Yapele, Vicky Jocelyne Ama-Moor

**Author notes:** Corresponding author: Eric Walter Pefura-Yone, Postal address: P.O Box-8340, Yaounde, Cameroon, Faculty of Medicine and Biomedical Sciences, University of Yaoundé 1, Yaoundé, Cameroon.

## Abstract

**Background:** Traditional diagnostic methods for Tuberculous pleural effusion (TPE) are often limited by their invasiveness, low sensitivity, and lack of accessibility. This study aims to develop and evaluate ML models for diagnosing TPE in African contexts, using readily available clinical and laboratory data.

**Methods:** A cross-sectional study carried out in Yaoundé, Cameroon (2018-2023), included patients with non-purulent exudative pleural effusion. Pleural fluid was analysed for total protein, lactate dehydrogenase (LDH), glucose, C-reactive protein (CRP) and cytology. TPE diagnosis relied on detection of tuberculous bacilli or tuberculous granuloma . Five ML models namely Random Forest (RF), XGBOOST, Logistic Regression (LR), Support Vector Machine (SVM), and Multilayer Perceptron (MLP) were tested using binary classification (TPE vs non-TPE) in Python software. The performance of models was evaluated using the area under the receiver operating characteristic curve (AUC), F1 score, accuracy, sensitivity, and precision.

**Results:** Of the 302 participants included, 175 (57.9%) were male and their median age (interquartile range) was 46 (34-61) years. Overall, 58.9% of participants had TPE, 15.9% had pleural metastasis and 25.2% had other causes of pleural effusion. Patients with TPE were younger, more often male and had a higher prevalence of HIV-infection. They also had higher pleural protein and CRP levels. The RF model showed the best performance with an AUC of 0.846 and an F1 score of 0.811 in the testing sample. Sensitivity was higher for MLP (0,944) and precision was higher for LR (0.806). Key predictors identified by the RF model were pleural CRP levels, age, pleural LDH levels, body mass index, and pleural protein levels.

**Conclusion:** The RF model had the best performance. MLP and LR had the best sensitivity and precision respectively. The models can be used to improve diagnosis of TPE in Africa settings.

**KEY MESSAGES:** *What is already known on this topic:* Tuberculous pleural effusion (TPE) diagnosis is invasive and lacks sensitivity in resource-limited African settings, with limited ML models available.

*What this study adds:* This study developed and evaluated five ML models for TPE diagnosis using clinical and pleural fluid data from Africa setting. The Random Forest model outperformed others in diagnosing TPE, using pleural CRP levels, age, pleural LDH levels, body mass index, and pleural protein levels as key predictors.

*How this study might affect research, practice, or policy:* Machine learning models can enhance TPE diagnosis in Africa using accessible and reliable biomarkers.

## INTRODUCTION

Tuberculous pleural effusion (TPE) represents a major public health concern in Africa, a region of the world where the prevalence of tuberculosis (TB) remains high[1–4]. In 2022, nearly 2.5 million people were living with TB in the African region, accounting for over 25% of new cases worldwide[5]. This burden is exacerbated by socio-economic inequalities, lack of adequate health infrastructure and the high prevalence of HIV infection[6,7]. Diagnosis of pleural tuberculosis in resource-limited regions is particularly difficult. Several factors contribute to these difficulties, including the invasive diagnostic procedures (pleural biopsy) required to obtain pleural samples, and the paucibacillary nature of pleural fluid, which gives poor results with traditional diagnostic methods such as direct microscopy and culture. These methods are often inaccessible or time-consuming[1]. Although new diagnostic tools such as GeneXpert have improved sensitivity, their high cost and limited availability in rural and low-resource areas still hamper diagnostic efficiency[8]. This scenario underscores the urgent need for innovative diagnostic approaches that enhance accuracy and accessibility for pleural TB in African contexts.

Machine learning (ML), a branch of artificial intelligence, is increasingly used in the healthcare field. It offers the possibility of analyzing large quantities of data and developing predictive models that improve the diagnosis, treatment and prognosis of diseases. In the field of tuberculosis diagnosis, ML algorithms have already shown their worth by improving the interpretation of chest X-rays, predicting drug resistance and optimizing diagnostic processes[9,10]. For TPE in particular, previous studies have begun to explore the use of ML to analyze clinical and biological data, integrating variables such as symptoms, biochemical markers and pleural fluid densitometric results[11–13]. These models hold promise for complementing the limitations of traditional diagnostic tools, particularly in resource-limited regions.

Despite these advances, several important gaps remain. Most existing studies on ML and TB focus primarily on pulmonary TB, neglecting extrapulmonary forms such as pleural TB[10]. In addition, many models have been developed in medium-to high-resource settings, raising questions about their applicability to African populations, where comorbidities and diagnostic constraints are different. This study aims to fill these gaps by developing and evaluating machine learning models specifically designed to diagnose TPE using data from African settings.

## METHODS

### Settings and participants

This cross-sectional study was conducted in the pulmonology departments of the Yaoundé Jamot Hospital (YJH) and the Polymère Private Clinic in Yaoundé, Cameroon from 1^st^ January 2018 to 31^st^ July 2023. The YJH is a teaching hospital that serves as a referral centre for respiratory diseases in Yaoundé and the surrounding area, while the Polymère Clinic is a specialist pneumology centre. The study involved participants aged 15 and over who presented with exudative, non-purulent pleural effusion. Patients were excluded if they did not have a pleural CRP assay or if they refused to take part in the study.

### Patient and Public Involvement Statement

Patients and/or the public were not involved in the design, or conduct, or reporting, or dissemination plans of this research.

### Data collection

Data collection involved face-to-face interviews, medical record reviews, clinical and paraclinical examinations. Demographic, clinical, haematological, pleural effusion and TB data were obtained. Demographic data included sex and age of participants. The clinical characteristics of the patients recorded were: presence or absence of respiratory symptoms (cough, dyspnoea, chest pain) and general symptoms (fever, hypersudation, weight loss), duration of symptoms, history of cancer or HIV infection, and body mass index (BMI). Haematological data included white blood cell count and haemoglobin level. The results of routine cytological (neutrophil count, presence of malignant cells) and biochemical analyses of pleural fluid, including total protein, lactate dehydrogenase (LDH) and glucose levels, were extracted from the participants’ files. The biochemical analysis of the pleural fluid was completed by measuring C-reactive protein. All CRP measurements were performed exclusively at the Polymère clinic laboratory. The method used to measure CRP levels was nephelometry, a light-scattering technique for determining protein concentrations (Genrui Biotech Inc., Shenzhen, China). The process involves introducing an anti-CRP antibody into the pleural fluid sample, which is contained in a specialized kit. The antibody reacts with the CRP in the sample, forming immune complexes that precipitate into tiny particles. The nephelometer measures the intensity of light scattered by these particles, which is directly proportional to the CRP concentration in the pleural fluid, enabling precise quantification.

Pleural cytology, closed pleural biopsy and the detection of tuberculous bacilli in sputum were used to investigate the causes of exudative pleurisy. The diagnosis of tuberculous pleural effusion was made either when tuberculous bacilli were detected by microscopic, culture or molecular (Xpert) methods in the pleural fluid, pleural biopsy specimen or sputum, or when a tuberculous granuloma was detected by histopathological analysis of pleural specimens. Subjects with the presence of malignant cells in the pleural fluid or on pleural biopsy specimens were classified as having pleural cancer metastasis. Other causes of non-purulent exudative pleurisy were investigated by appropriate paraclinical methods. Ethical clearance was obtained from the Faculty of Medicine and Biomedical Sciences, University of Yaounde 1. Consent or assent from the guardians of all participants was also obtained.

### Data management

Data were analyzed using Python version 3.12.4. Categorical variables were summarized using counts and percentages, while quantitative variables were presented as medians and interquartile ranges due to their non-Gaussian distribution. The chi-square test or Fisher’s exact test was used to compare categorical variables. The Mann-Whitney U test was applied for comparisons between two groups of quantitative variables. We selected the variables associated with TPE with p < 0.10 as independent variables for the development and evaluation of ML models. We used binary classification (TPE vs non-TPE) to develop and evaluate five ML models namely Random Forest (RF), XGBOOST, Logistic Regression (LR), Support Vector Machine (SVM), and Artificial Neural Network (i.e: multilayer perceptron, MLP). The scikit-learn package version 1.4.2 was used for the development of all models except for the XGBOOST for which we used the xgboost package version 2.1.3. For model development and evaluation, the sample was divided into a training set (80%) and test set (20%). Missing values in quantitative variables were replaced by their medians. Quantitative variables were standardized and qualitative variables were numerically encoded. The same data preparation procedures were applied in the training sample as in the test sample. The training sample was used for model training, hyperparameter search using randomized hyperparameter search, and cross-validation. The test sample was used exclusively for final model evaluation. The area under the Receiver Operating Characteristic(ROC) curve (AUC) was the main optimized metric. Other metrics calculated were F1 score, accuracy, sensitivity, and precision. The features’ importance was measured when possible. The codes used will be available as supplementary files.

## RESULTS

### General Characteristics of participants

The study included a total of 302 participants (after exclusion of 44 participants with incomplete data on key variables), with a median age of 46 years [interquartile range (IQR): 34–61 years]. Of the participants, 57.9% were male, and 42.1% were female. Most participants (96.0%) reported experiencing a cough, while 94.7% had dyspnea. Chest pain was reported by 82.5% of individuals, and 39.7% had symptoms lasting one month or less. Excessive sweating was reported by 49.7% of individuals, and 23.8% were HIV-positive. Among the participants, 5.0% had a history of cancer. The median protein levels in pleural fluid was 42.8 g/L (IQR: 36.6–53.0 g/L), while the median CRP levels in pleural fluid was 42.9 mg/L (IQR: 24.9–62.8 mg/L). The general characteristics of participants are summarized in Table 1.

**Table 1:** Baseline clinical characteristics and pleural fluid analysis

| Characteristics | Overall<br>n = 302 (%) | TPE<br>n = 179 (%) | non-TPE<br>n = 123 (%) | p |
| --- | --- | --- | --- | --- |
| Age (median [IQR]), years | 46.0 [34.0, 61.0] | 41.0 [30.0, 52.0] | 54.00 [40.50, 67.00] | 0.001 |
| Male sex(%) | 175 (57.9) | 118 (65.9) | 57 (46.3) | 0.001 |
| Cough | 290 (96.0) | 176 (98.3) | 114 (92.7) | 0.030 |
| Dyspnea | 286 (94.7) | 171 (95.5) | 115 (93.5) | 0.607 |
| Chest pain | 249 (82.5) | 150 (83.8) | 99 (80.5) | 0.485 |
| Duration, months |  |  |  |  |
| ≤ 1 | 120 (39.7) | 75 (41.9) | 45 (36.6) | 0.020 |
| ]1, 2] | 94 (31.1) | 56 (31.3) | 38 (30.9) |  |
| ]2, 3] | 60 (19.9) | 39 (21.8) | 21 (17.1) |  |
| > 3 | 28 (9.3) | 9 (5) | 19 (15.4) |  |
| Asthenia | 272 (90.1) | 163 (91.1) | 109 (88.6) | 0.616 |
| Anorexia | 263 (87.1) | 159 (88.8) | 104 (84.6) | 0.361 |
| Weight loss | 277 (91.7) | 170 (95.0) | 107 (87.0) | 0.024 |
| Fever | 233 (77.2) | 159 (88.8) | 74 (60.2) | <0.001 |
| Sweating | 150 (49.7) | 98 (54.7) | 52 (42.3) | 0.044 |
| HIV infection | 72 (23.8) | 55 (30.7) | 17 (13.8) | 0.001 |
| Cancer | 15 ( 5.0) | 0 ( 0.0) | 15 (12.2) | <0.001 |
| BMI (median [IQR]), kg/m<sup>2</sup> | 21.3 [19.6, 23.7] | 20.8 [19.3, 22.36] | 22.3 [20.5, 25.1] | <0.001 |
| Blood Haemoglobin (median [IQR]), g/dL (n = 140) | 10.0 [8.1, 11.9] | 10.2 [8.5, 11.8] | 9.6 [7.7, 11.9] | 0.458 |
| Blood WBC count (median [IQR]), cells/uL (n = 139) | 7000.0 [5050.0, 9900.0] | 6900.0 [5200.0, 8945.0] | 7600.0 [5000.0, 11225.0] | 0.533 |
| Pleural proteins (median [IQR]), g/L | 42.8 [36.6, 53.0] | 47.6 [38.0, 55.2] | 39.6 [34.8, 45.1] | <0.001 |
| Pleural CRP (median [IQR]), g/L | 42.9 [24.9, 62.8] | 50.2 [30.7, 75.5] | 30.2 [16.4, 51.0] | <0.001 |
| Pleural LDH (median [IQR]), UI/L | 645.0 [45.50, 982.2] | 709.0 [539.0, 1063.0] | 540.0 [387.0, 806.0] | <0.001 |
| Pleural glucose (median [IQR]), g/L | 0.86 [0.67, 1.04] | 0.86 [0.68, 1.02] | 0.87 [0.64, 1.06] | 0.677 |
| Pleural PMN (median [IQR]), %, (n = 190) | 15.0 [5.0, 27.0] | 13.0 [3.5, 25.2] | 20.0 [9.2, 29.7] | 0.024 |

TPE, tuberculous pleural effusion; IQR, interquartile range; HIV, human immuno-deficiency virus; BMI, body mass index; WBC, white blood cells count; CRP, C-reactive protein; LDH, lactate dehydrogenase; PMN, polymorphonuclear cells

### Prevalence and predictors of tuberculous pleural effusion

Overall, 178 (58.9%) participants had TPE and 124 (41.1%) had another cause of non-purulent exudative pleurisy, including 48 (15.9%) pleural metastases. There were notable differences between participants with TPE and those without TPE. Patients with TPE were younger, with a median age of 41 years compared with 54 years for patients without TPE (p < 0.001). Male subjects were more represented in the TPE group (65.9% vs 46.3%, p = 0.001). Patients with TPE were more likely to present with symptoms such as cough (98.3% vs 92.7%, p = 0.017), fever (88.8% vs 60.2%, p < 0.001), and weight loss (95.0% vs 87.0%, p = 0.024). The prevalence of HIV infection was significantly higher in TPE patients (30.7%) than in non-TPE patients (13.8%, p = 0.001). Biochemical analysis showed that TPE patients had higher median pleural fluid protein levels (47.6 g/L vs 39.6 g/L, p < 0.001) and higher median pleural fluid CRP levels (50.2 mg/L vs 30.2 mg/L). These results are detailed in Table 1.

### Performance of models in cross-validation for TPE diagnosis

The learning curves presented in Figure 1 show the evolution of the performance of each model measured using AUC as a function of the training data size. The RF and XGBoost models showed consistently high training scores, with plateauing cross-validation scores. LR, SVM, and MLP showed decreasing training scores and increasing cross-validation scores, converging towards larger training sizes. All models showed a plateau, suggesting limited benefit from more data.

**Figure 1:**
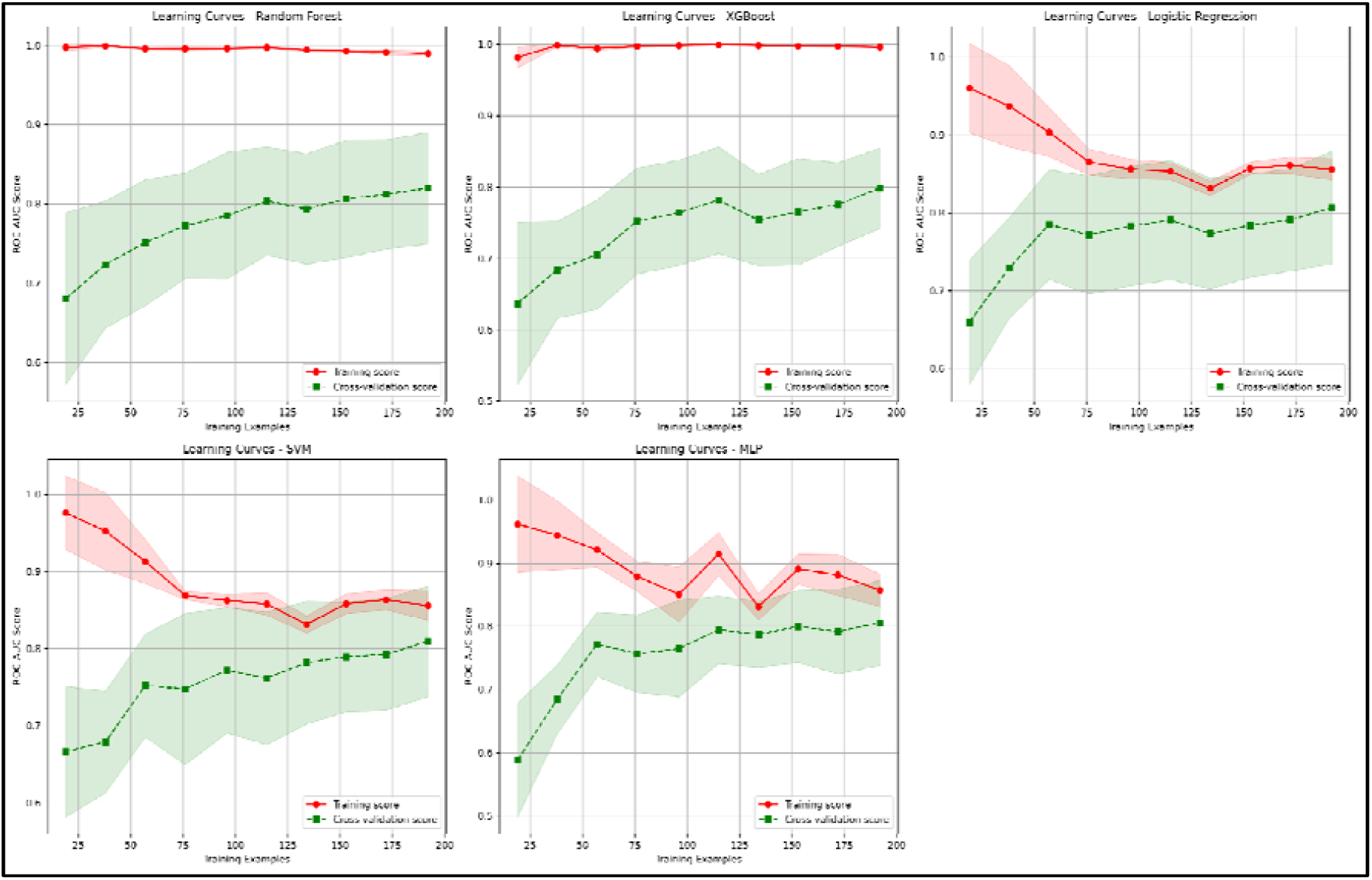
Learning curves of machine learning models. ROC-AUC, area under the receiver operating characteristic curve

During cross-validation, the RF classifier achieved the highest average AUC of 0.814 (±0.080), demonstrating good predictive performance. LR followed closely with an average AUC of 0.798 (±0.074), while XGBoost achieved 0.775 (±0.054), showing consistent but slightly lower predictive ability. For the SVM the average AUC was 0.770 (±0.076). The MLP recorded the lowest average AUC of 0.752 (±0.034), indicating average but less competitive performance compared to other models.

### Final comparison of models on testing set for TPE diagnosis

The final evaluation of the models on the test sample is presented in Table 2 and Figure 2. The RF model obtained the highest AUC (0.846) and F1 score (0.811), as well as high recall (0.833), precision (0.789), and accuracy (0.770), making it the best performing model. LR provided competitive performance with an AUC of 0.838, precision (0.806), recall (0.806), and F1 score of 0.806, as well as a more favorable bias-variance tradeoff. The worst performing model was the MLP with the lowest accuracy (0.623) and AUC of 0.800. On the other hand, the MLP had the highest sensitivity (0.944).

**Table 2:** Performance metrics in testing set

| **Models** | **Accuracy** | **Precision** | **Recall** | **F1-score** | **AUC** |
| --- | --- | --- | --- | --- | --- |
| Random Forest | 0.770 | 0.789 | 0.833 | 0.811 | 0.846 |
| XGBoost | 0.721 | 0.771 | 0.750 | 0.761 | 0.842 |
| Logistic<br>Regression | 0.770 | 0.806 | 0.806 | 0.806 | 0.838 |
| SVM | 0.738 | 0.778 | 0.778 | 0.778 | 0.840 |
| MLP | 0.623 | 0.618 | 0.944 | 0.747 | 0.800 |

SVM, support vector machine; MLP, multilayer perceptron; AUC, area under the receiver operating characteristic curve

**Figure 2:**
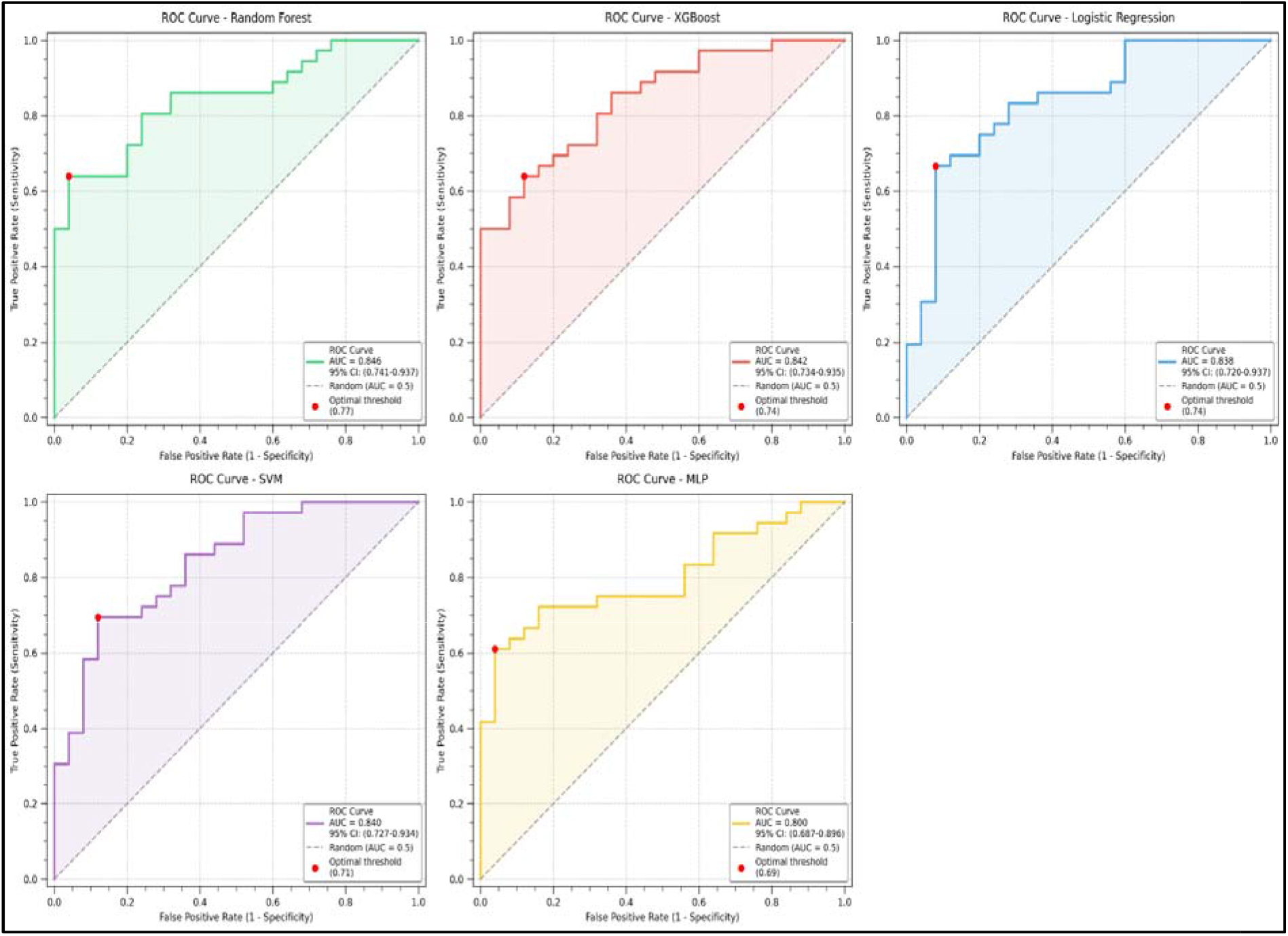
Area Under the Receiver Operating Characteristic Curve (AUC-ROC) in the testing set. ROC-AUC, area under the receiver operating characteristic curve, SVM, support vector machine; MLP, multilayer perceptron

**Figure 3:**
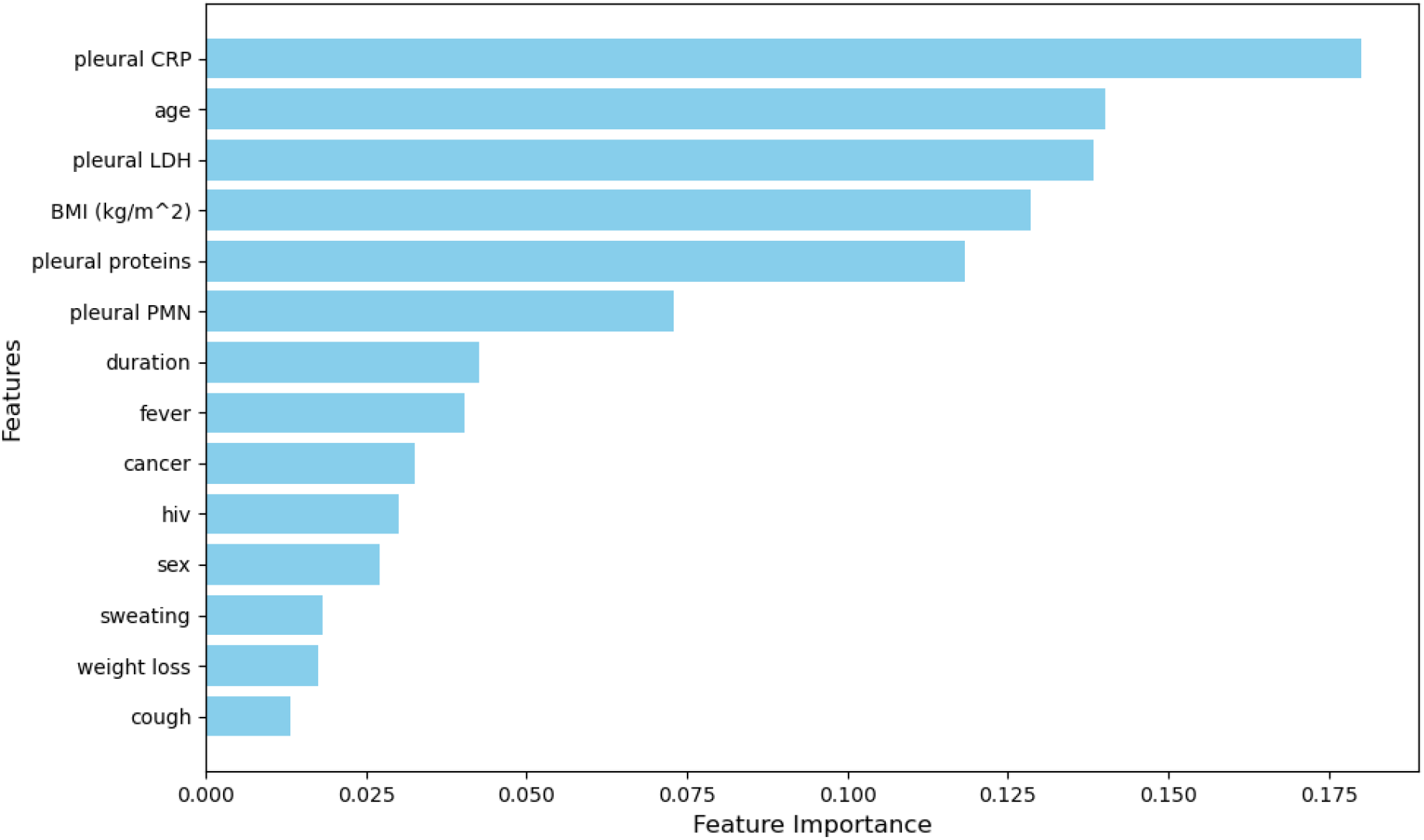
Random forest features importance

## DISCUSSION

This study developed and evaluated machine learning models for the diagnosis of tuberculous pleural effusion (TPE) in patients with non-purulent exudative pleural effusion in African settings with high prevalence of TB. It is a reliable and accessible diagnostic tool that could easily complement traditional methods, which have their limitations in resource-limited settings. The models used readily available variables such as patient characteristics, symptoms, and pleural fluid analysis to predict the possibility of pleural TB. Among the models, the performance of the RF model was the best in this study, with an AUC of 0.846, F1 score of 0.811, recall of 0.833 and precision of 0.789. The main five predictors were pleural CRP levels, age, pleural LDH levels, BMI, and pleural protein levels, and they account for 70% of the predictive power of the RF model.

Identification of key predictors for TPE is a very important part of improving diagnostic precision and guiding clinical decisions[14–16]. This combination of readily available clinical and biochemical markers, therefore, forms the basis of our analysis, with pleural fluid CRP emerging as a very strong predictor. Studies show that the CRP levels in TPE are significantly higher when compared to other types, especially malignant pleural effusions[17,18]. High CRP in the pleural fluid reflects the significant inflammatory response that is characteristic of tuberculous infection. In one study, high CRP levels (≥ 50 mg/l) have a high specificity for tuberculosis (95%), and low levels (<30 mg/l) have a high sensitivity (95%) for excluding disease[18]. Wu et al. developed a scoring model to differentiate TPE from non-TPE based on six variables: age ≤46 years, male gender, absence of cancer, positive T cell-SPOT, ADA ≥24.5 U/L, and CRP ≥52.8 mg/L. The model demonstrated high sensitivity (93.7%), specificity (96.8%), and accuracy (99.2%). Validation confirmed its robust performance, achieving high sensitivity (92.9%), specificity (93.3%), and accuracy (93.1%)[14].

Age is also an important factor associated with TPE. In most studies, younger age is associated with TPE[11,12,14,19]. This observation may indicate a higher prevalence of other causes of exudative pleural effusion, such as pleural metastases, in older individuals. Pleural fluid LDH levels also showed good predictive capability in diagnosing TPE. This is in agreement with the general literature, which puts LDH forward as a marker of cellular damage and inflammation, both being salient features in TPE. High levels of LDH in pleural fluid indicate active cellular turnover and tissue damage within the pleural space and are indicative of an underlying infectious process. The combination of LDH with other markers, such as adenosine deaminase (ADA), is also indicated to be a strong predictor of TPE[15,19]. The other important aspect to consider is the pleural protein content. TPEs present as exudative effusions, with a higher protein content. This increased protein level reflects the permeability of pleural vessels due to inflammation and, therefore, suggests a diagnosis of TPE in association with clinical and other laboratory findings.

Although individual markers give a very good overview of the disease, machine learning analyses and scoring systems suggest that the synergistic effect of these predictors is very useful. In general, models including these factors tend to outperform those using a single marker, reinforcing the importance of a comprehensive multiparametric approach to the accurate diagnosis of TPE[12,15]. Therefore, the models which integrate clinical information on symptoms and patients characteristics with laboratory results, can provide a more sensitive and reliable predictive capability for TPE.

Very few data have been published on the use of ML models to help diagnose tuberculous pleurisy. Comparison of the performance of ML models in the diagnosis of TPE is made difficult by the variation in the parameters included in the models. Most of the few published models have included pleural ADA level as one of the independent variables. To date, no published ML model has included pleural CRP levels. In our study, the RF model showed the highest performance, although the AUC of the other models tested also suggested good performance of these models. The first published (2013) model was based on artificial neural networks and the reported performance was based on cross-validation without independent sample evaluation. In this study, the pre-test sensitivity and specificity of the neural network were 94.5% and 91% respectively during cross-validation. The other studies tested various ML models including logistic regression, support vector machine, k-nearest neighbour, decision tree, random forest and boosting gradient machine [20].

In the study by Ren et al. carried out in China, RF was the best performing model with an AUC of 0.965 and the most important predictors were pleural ADA level, pleural carcinoembryonic antigen (CEA Ag) level and age. On the other hand, in the study by Liu et al (also carried out in China), the best performing model for discriminating between TPE and non-TPE was the SVM with an AUC of 0.914. The three most important predictors in the study by Liu et al. were pleural effusion ADA, pleural effusion CEA Ag, and serum CYFRA 21-1[11]. In another study carried out in a country with a low prevalence of tuberculosis (Spain), SVM was the best performing model with an AUC of 0.98 in the testing set. In this study, pleural ADA, age and pleural lymphocyte count > 50% were the important predictors [12].

The main limitation of our study is the non-measurement of pleural ADA in our participants. Indeed, measurement of pleural fluid ADA has shown a uniformly high diagnostic performance for TPE in five major consecutive meta-analyses with a mean sensitivity and specificity of 92-93% and 90-92% respectively for a diagnostic threshold of >40 U/l. Measurement of pleural ADA in our patients would therefore improve the performance of our model. Pleural ADA measurement is not routinely performed in many hospitals in sub-Saharan Africa, so the models developed in our study are based on parameters routinely measured in this context. Pleural CRP is a marker that is available even in areas with limited resources, and its inclusion in the models has improved their performance.

## CONCLUSION

This study successfully developed and evaluated machine learning models for diagnosing TPE in African settings, using readily available variables. The RF model demonstrated the best performance, achieving an AUC of 0.846 and an F1-score of 0.81. MLP and LR had the best sensitivity (0.944) and accuracy (0.806) respectively. The main predictors were pleural CRP levels, age, pleural LDH levels, BMI and pleural protein levels. The models offer a reliable and accessible diagnostic tool to supplement the limitations of traditional methods in resource-limited settings.

## Data Availability

All data produced in the present study are available upon reasonable request to the authors

## Contributors

EWPY and VJAM conceived the study, supervised data collection, co-analysed the data and drafted of the manuscript; ADB, LMEM, JB, MCE, CMY collected data, contributed to data analysis and critically revised the manuscript; MM, MENK, VPM, AK, AWN, DA, ALNN, PLTN, MJNE contributed to data analysis and critically revised the manuscript; All authors approved the final version of the manuscript.

## Data availability statement

Data are available upon reasonable request.

## Ethics statements

This study involves human participants. The research was approved by the institutional ethical committee of the Faculty of Medicine and Biomedical Sciences, University of Yaoundé 1 (reference 0043/2018). Participants gave informed consent to participate in the study before taking part.

## Acknowledgements

The authors would like to thank the invaluable contributions by the study participants and data collection staff.

